# Design and Methods of the “Understanding pre-exposure prophylaxis modalities for HIV prevention in European communities” (PROTECT) Survey

**DOI:** 10.1101/2024.11.04.24316697

**Authors:** Johann Kolstee, Haoyi Wang, Hanne Zimmermann, Melanie Schroeder, Ama Appiah, Carolyn Brown, Ana Milinkovic, Kai J. Jonas

**Affiliations:** Maastricht University, Faculty of Psychology and Neuroscience - Maastricht, Netherlands; ViiV Healthcare Ltd, Brentford, UK; ViiV Healthcare Ltd, Durham, NC USA; Chelsea and Westminster Hospital, London, UK

## Abstract

**Background:** Despite the increasing availability of oral PrEP in Europe, gaps remain in the coverage of HIV prevention strategies at a population level. Long-acting injectable PrEP (LA PrEP) has the potential to help provide greater HIV prevention coverage to increasing proportions of men who have sex with men (MSM) and trans people, communities still disproportionately affected by HIV.

**Methods:** An online cross-sectional survey was conducted across 20 European countries from October 2023 to April 2024. The survey was translated into 22 local and migrant languages. The main aim of the survey was to understand which MSM and trans* people in Europe are interested and intend to use LA-PrEP if it becomes available.

**Results:** A total of 15,458 MSM and trans* individuals participated in the survey. Participants were recruited mainly from gay dating apps and through a social media campaign. Overall, the sample was sexually active (96.4%), engaged in condomless anal intercourse in the past 6 months (83.4%), had more than 10 partners in the past 6 months (57.6%) and was PrEP naïve (51.4%).

**Conclusions:** MSM and trans people in Europe can be engaged effectively to participate in research about LA PrEP.

## INTRODUCTION

HIV remains a serious health threat in Europe (1-4). Men who have sex with men (MSM) and trans* people continue to bare a disproportionately large burden of new HIV infections in Europe (3-5). In countries within the European Union and European Economic Area in 2022 there were 22,995 new HIV infections reported, with 33.3% being reported among MSM (4). Despite being a region in which universal health care is widely available, HIV prevention efforts, and in particular access to new biomedical advances such as oral HIV pre-exposure prophylaxis (PrEP), have been patchy (6, 7) or have been placed rather late within national healthcare policies (e.g. Italy in 2023) (8) .

Increasing proportions of new infections are being reported among heterosexual individuals and migrant populations, which is driving the epidemic in new directions (4, 9, 10). Divergence exists between western and eastern Europe, with the epidemic in eastern Europe representing the majority of new infections on the continent in 2022 (4). Prevention efforts in eastern Europe have not been as consistent or well-funded, and the war in Ukraine has created additional challenges for health promotion (11, 12).

Despite these structural and healthcare hurdles, awareness of oral PrEP has grown, as have positive attitudes towards taking it, among MSM in Europe over the past decade (13). With limits on how pharmaceutical companies can market new medications to consumers, it has fallen on health systems and affected communities themselves to spread the word about oral PrEP. Access to oral PrEP has taken several years to expand in Europe, with some countries still not making it widely available today to communities most affected by HIV (14). Despite this, interest in PrEP exists among MSM in many European countries (15-17). Although awareness, interest and access to PrEP in Europe has expanded, PrEP uptake has not yet met the UN HIV prevention 2025 targets of PrEP use among MSM of 50% (18, 19). Increased PrEP uptake, along with condom use and the use of treatment as prevention (TasP) have the potential to significantly reduce HIV infections (20).

The development of alternative modalities to deliver PrEP provides opportunities to increase population-level PrEP coverage among people at risk of HIV (3), and have been shown to be of interest to populations such as MSM (21-23). In 2023, the European Medicines Agency approved Cabotegravir, a long-acting injectable form of HIV PrEP (LA-PrEP), for use in Europe (24). In light of this new HIV prevention tool becoming available, it is important to understand who may benefit from this innovation, if access becomes widespread in Europe. It is this question that drove the development of the, “*Understanding pre-exposure prophylaxis modalities for HIV prevention in European communities”,* (PROTECT) study.

Large surveys of European MSM have been running, since the advent of the HIV epidemic, to understand HIV risk and prevention behaviors (25-27). This tradition of research has adopted online data collection methods over the past few decades (28). This mirrored the movement online of many MSM seeking sexual partners and creating communities (29, 30). From 2010 to 2017 the European men who have sex with men internet survey (EMIS) was regularly engaged MSM across 50 countries in Europe about HIV and sexual health (27). The PROTECT survey exists within and has been informed by this broader research tradition.

In this paper we demonstrate how the PROTECT survey was developed and implemented. We provide a breakdown of who participated and from where they participated. The sample characteristics are also described in this paper. The aim of the PROTECT study was to understand which MSM and trans* individuals in Europe are interested and intend to use LA-PrEP if it becomes available.

## METHODS

The PROTECT survey sought to understand who in Europe is interested in, and has the intention to use, LA-PrEP, and in particular which MSM and trans* individuals. The team at Maastricht University leading the project leveraged knowledge built through previous pan-European work on this population and topic. The PROTECT study protocol was reviewed and approved by the Ethics Review Committee Psychology and Neuroscience (ERCPN) at Maastricht University (OZL_262_08_01_2023_S21).

### Target population

MSM and trans* individuals 18 years of age or older and living in the following twenty countries were eligible to participate: Austria, Belgium, Cyprus, Czech Republic, Denmark, Finland, France, Germany, Greece, Ireland, Italy, Luxemburg, Netherlands, Norway, Poland, Portugal, Spain, Sweden, Switzerland, and the United Kingdom. The research team itself was comprised mainly of individuals from these target populations. Furthermore, individuals from these target populations were also selected to undertake various translations of the survey, to support survey testing and to help develop the campaign materials and promotional approach described later. The responses of people living with HIV were also included to explore their attitudes towards their HIV negative partner’s potential use of LA-PrEP and their attitudes towards long-acting anti-retroviral treatment (LA-ART).

### Questionnaire development

An online survey was developed to gather data from participants. Qualtrics XM (R) software was used to collect data. The questionnaire was developed with the primary research aim in mind, to understand who in Europe is interested in, and has the intention to use, LA-PrEP. Once a draft questionnaire was developed, it was shared with a panel of community representatives for review and feedback. Individuals and organizations representing people living with HIV and trans* individuals were engaged as were experts from non-European backgrounds. The guidance provided by these experts and organizations was incorporated and helped tailoring the survey.

Extensive survey testing was undertaken to ensure that the question logics for each section of the survey flowed correctly. This survey testing was conducted on the English version of the survey prior to it being translated into the twenty-one other languages (Arabic, Czech, Danish, Dutch, Finnish, French, German, Greek, Hebrew, Italian, Mandarin, Norwegian, Polish, Portuguese, Russian, Spanish, Swahili, Swedish, Turkish, Ukrainian, and Urdu). These languages represented the local language of each country in which the survey was launched, and select migrant languages relevant to the countries surveyed. Subsequently, all the translated versions of the survey were also tested to ensure the correct sequence and flow of questions throughout. Care was taken to ensure that key terms were translated correctly and where possible community-based translators with expertise in health related, and HIV related, translation experience were selected. Participants in every location could choose to take the survey in any of the 22 languages.

### Measures

The questionnaire contained the following sections: Socio-demographics; sexual behavior; HIV and STI testing and status; mental health; psychological variables; knowledge, attitudes and beliefs towards HIV, PrEP and UVL; experiences with oral PrEP; PrEP discontinuation; LA-PrEP interest and intention; experiences of people living with HIV, see table 1 below for more detailed information. The total number of possible questions received by participants varied based on their characteristics. Current PrEP users had 93 questions, former PrEP users 91, PrEP naïve people 83, and people living with HIV had 73 questions in the survey. Some question sets were based on questions from EMIS-2017, such as questions about sexual behavior, sexual self-efficacy, depression/anxiety and PrEP use (27). Others were based on previously validated scales, such as the general self-efficacy scale, the openness to new experiences scale and the brief version of the Big Five Personality Inventory (31-33).

**Table 1:** Composition of PROTECT questionnaire

| **Section** | **Questions** |
| --- | --- |
| **Socio-demographics** | Country, region, gender identity, sex assigned at birth, sexual orientation, birth year, size of location of residence, housing situation, birth county self/mother/father, language most commonly spoken at home, educational level, current occupation, gross annual income, comfort on current income scale, connection to LGBTQI+ communities scale, current relationship status |
| **Sexual behavior** | Number of sex partners in last 6 months, anal sex in the last 6 months, position taken most often in anal sex, condomless anal sex in the last 6 months, vaginal sex in the last 6 months, condomless vaginal sex in the last 6 months, drug use for sex in last 6 months, which drugs, giving payment for sex, receiving payment for sex |
| **HIV and STI testing and status** | HIV status, HIV testing frequency, STI testing frequency, STI diagnosis ever, which STIs diagnosed, vaccinations received |
| **Mental health** | Depression/anxiety (PHQ4), psychosis, self-harm, attention deficit disorder |
| **Other psychological scales** | Comfort with current sexual orientation and gender identity, homo-negativity, self-efficacy, openness to experiences, sexual self-efficacy, satisfaction with sex life, satisfaction with life in general, Big Five Personality Inventory brief version |
| **Knowledge, attitudes and beliefs towards HIV, PrEP and undetectable viral load (UVL)** | Likelihood of acquiring HIV, worry about acquiring HIV, communication about HIV prevention with partners, PrEP awareness, PrEP use current, PrEP use past, attitudes towards PrEP, impact of PrEP use, attitudes towards HIV treatment, awareness of LA PrEP, LA PrEP use |
| **Former oral PrEP users** | Previous access to oral PrEP, ideal oral PrEP access options, reasons for stopping oral PrEP, alternatives to oral PrEP use, past PrEP type, past PrEP regime, duration of previous PrEP use, preferred way to take PrEP, use of post exposure prophylaxis for HIV (PEP) |
| **Current oral PrEP users** | Current oral PrEP regime, current PrEP access, ideal PrEP access, duration of current PrEP use, likelihood of ongoing PrEP use, on demand pre sex dosing, any missing pills for daily users, shortening of pre sex dosing for on demand, frequency of forgetting to take pills post sex for on demand, kidney test in past 6 months, use of PEP |
| **Oral PrEP access** | Unsuccessful attempts to acquire oral PrEP, offers of PrEP access from healthcare providers, reasons for why healthcare providers offered oral PrEP, ease to obtain oral PrEP currently |
| **Oral PrEP access barriers** | Potential barriers experienced in accessing oral PrEP |
| **LA PrEP interest and intention** | Interest in taking LA PrEP, likelihood of starting LA PrEP, likelihood of combining LA PrEP with other PrEP modalities, potential perceived benefits of LA PrEP use, willingness to pay for LA PrEP, willingness for particular price points to pay for LA PrEP, concerns with LA PrEP use, presence of gluteal implants, willingness to remove gluteal implants for potential LA PrEP use, preference for PrEP modalities if planning to get gluteal implants, worries about LA PrEP and hormone use interactions, worries about LA PrEP impacting bone mineral density, preferred frequency of LA PrEP injection, likelihood of attendance at appointments every 2 months for injections, ideal LA PrEP provider, word cloud what does LA PrEP mean to you |
| **LA PrEP mixed use and informal procurement** | Likelihood of combining LA PrEP and oral PrEP regimes, likelihood of using LA PrEP at certain seasons of risk during the year, likelihood of travelling and using LA PrEP in other countries were available and affordable, potential actions if LA PrEP was available and then made not available in your country |
| **PLHIV – HIV testing and status** | Year of HIV diagnosis, treatment status, year treatment commenced, undetectable viral load, worry about transmitting HIV to sex partners |
| **PLHIV – LA PrEP interest and intention to use** | Status of current sex partner if in relationship, PrEP use in partner, preference for partner to take LA PrEP over oral PrEP, willingness to contribute to paying for partners LA PrEP, price willing to help partner pay for LA PrEP, comfort with missing own treatment if partner on LA PrEP or oral PrEP |
| **PLHIV – LA PrEP & LA ART Attitudes and Beliefs** | Attitudes towards LA PrEP effectiveness and interest to use LA ART |

#### The Patient Health Questionnaire for Depression and Anxiety (PHQ-4) scale

Anxiety and depression was assessed using the PHQ-4 scale (34). This scale has 4 items that participants responded to by selecting a response on a 4-point Likert scale from 1 (not at all) to 4 (nearly every day). For example, participants were asked to rate the following item, *“over the last 2 weeks, how often have you been bothered by the following problems (item: Feeling nervous, anxious or on edge)?”.* The internal reliability for this scale was very good (α = 0.87).

#### General self-efficacy

General self-efficacy was assessed using a 3-item scale that participants responded to by selecting a response on a 5-point Likert scale from 1 (strongly disagree) to 5 (strongly agree) (31). For example, participants were asked to rate the following item, “*I can always manage to solve difficult problems if I try hard enough”.* The internal reliability for this scale was good (α = 0.79).

#### Sexual self-efficacy

Sexual self-efficacy was assessed using a 2-item scale that participants responded to by selecting a response on a 5-point Likert scale from 1 (strongly disagree) to 5 (strongly agree). For example, participants were asked to rate the following item, “*The sex I have is always as safe as I want it to be”.* The correlation of the two items was medium (*r* = 0.53).

#### Openness to new experiences

Openness to new experiences was assessed using a 9-item scale that participants responded to by selecting a response on a 5-point Likert scale from 1 (strongly disagree) to 5 (strongly agree) (31). For example, participants were asked to rate the following item, “*I don’t like trying new things and would rather stick with what I know”.* The internal reliability for this scale was weaker, but still acceptable (α = 0.68).

#### Brief version of the Big Five Personality Inventory

Personality was assessed using the brief version of the Big Five Personality Inventory scale (33). This scale has 10 items that participants responded to by selecting a response on a 5-point Likert scale from 1 (strongly disagree) to 5 (strongly agree). For example, participants were asked to rate the following item, *“I see myself as someone who is reserved”.* The correlation pattern showed mixed levels: extraversion items (*r* = 0.45), the two items related to agreeableness was *r* = 0.10, the two items related to conscientiousness was *r* = 0.20, the two items related to neuroticism was *r* = 0.47 and the two items related to openness was *r* = 0.21.

#### Internalised homonegativity

Internalised homonegativity was assessed using a 7-item scale that participants responded to by selecting a response on a 5-point Likert scale from 1 (strongly disagree) to 5 (strongly agree) (35). For example, participants were asked to rate the following item, “*I feel comfortable in gay bars”.* The Cronbach’s alpha for this scale was good (α = 0.78).

Separately, comfort with sexual orientation and gender identity were measured using a 3-item scale that participants responded to by selecting a response on a 5-point scale from 1 (strongly disagree) to 5 (strongly agree). The three items in this scale were: *“I feel comfortable with my sexual orientation”, “I feel comfortable with my gender identity”, “I feel comfortable with my sexual characteristics (e.g. current physical traits)”*. The consistency of this scale was good (α = 0.75).

#### Likelihood of acquiring HIV

Likelihood of acquiring HIV was assessed using a 1-item assessment that participants responded to by selecting a response on a 5-point Likert scale from 1 (extremely unlikely) to 5 (extremely likely). Participants were asked to rate the following item, “*How likely do you think you will become HIV positive?”*.

#### Worry about acquiring HIV

Worry about acquiring HIV was assessed using a 1-item assessment that participants responded to by selecting a response on a 5-point Likert scale from 1 (never) to 5 (always). Participants were asked to rate the following item, “*How often do you worry about getting HIV?”*.

#### Attitudes towards PrEP

Attitudes towards PrEP, among PrEP naïve participants, were assessed using a 12-item scale that participants responded to by selecting a response on a 5-point Likert scale from 1 (strongly disagree) to 5 (strongly agree) (36). For example, participants were asked to rate the following item, “*I would be willing to take PrEP to prevent getting HIV”.* The Cronbach’s alpha for this scale was good (α = 0.76).

Attitudes towards PrEP, among participants currently using PrEP, were assessed using a 4-item Likert scale that participants responded to by selecting a response on a 5-point Likert scale from 1 (strongly disagree) to 5 (strongly agree) (36). For example, participants were asked to rate the following item, “*I am less worried about getting HIV because of PrEP”*.

#### Attitudes towards HIV treatment scale

Attitudes towards HIV treatment, among all participants, were assessed using a 3-item Likert scale that participants responded to by selecting a response on a five point scale from 1 (strongly disagree) to 5 (strongly agree) (36). For example, participants were asked to rate the following item, “*An HIV positive person who is on HIV treatments is unlikely to transmit HIV”.* The Cronbach’s alpha for this scale was good (α = 0.70).

#### LA PrEP intention

Intention to use LA PrEP was assessed using a 1-item assessment that participants responded to by selecting a response on a 5-point Likert scale from 1 (Extremely unlikely) to 5 (Extremely likely). Participants were asked to rate the following item, “*How likely are you to start using LA PrEP every 2 months if it is available to you?”*.

#### LA PrEP mixed use

Intention to mix use of LA PrEP with other PrEP modalities was assessed using a 1-item assessment that participants responded to by selecting a response on a 5-point Likert scale from 1 (Extremely unlikely) to 5 (Extremely likely). Participants were asked to rate the following item, “*How likely would you be to use LA PrEP on demand, for example, having injections for the summer period and then going back to using oral PrEP or condoms during other seasons?”*.

#### Perceived Benefits of LA PrEP

The perceived benefits of LA PrEP were assessed using a 5-item scale that participants responded to by selecting a response on a 5-point Likert scale from 1 (strongly disagree) to 5 (strongly agree). For example, participants were asked to rate the following item, “*Taking LA PrEP would release me from thinking about taking a pill every day”.* The Cronbach’s alpha for this scale was good (α = 0.76).

### Promotion and recruitment

The PROTECT survey was promoted to MSM and trans* individuals online between October 2023 and April 2024. The campaign approach included four main components, a website, social media advertisements (Instagram and Facebook), dating app advertisements (Grindr and Hornet) and flyers. The study team engaged a communications agency to help develop branding, the website and marketing collateral for the survey’s promotion. People from the survey’s target population informed the selection of imagery and style. A set of images were selected for the campaign, primarily of people, and these were used across the twenty countries in which the survey was conducted. A unifying design concept created consistency across the marketing collateral. A slogan was included on each image used for social media and dating app advertisements and translated into select languages. The slogan was: *Inject PrEP to protect? Take part in a survey now!* Each post on social media had additional information about what the survey was about in the description space below promotional images. The survey website contained: more detailed information about the purpose of the research, news on the topic, the research team and links to organizations that could provide participants with more information about PrEP in their respective countries. All imagery, and the website and flyers, linked participants to the online survey where more information was provided. Consent was required before participants could proceed to complete the survey.

In addition to the promotion described above to MSM and trans* individuals, the PROTECT survey was also promoted to heterosexual populations in Europe. Data collection and analysis for that arm of the survey will be described in a separate paper. However, participants who identified as men who have sex with men and participated in the survey via the heterosexual promotion have been included in the sample described in this analysis.

### Analysis

In this analysis we have provided information about participation in the PROTECT survey across the whole sample and by country and by promotion source. We have analyzed the retention rate by reporting the number of participants who attempted to complete any questions in our survey and those who completed more than 95% of the survey. This later group was obtained by cleaning the data set to remove any responses that were deemed to be erroneous and that were less than 95% complete. To prevent fraudulent responses open and logic questions were included to identify internet bots. The 95% completion threshold did not affect the main content questions of the survey. All questions in the PROTECT survey were designed so that for a participant to move forward to the next question, they had to answer every previous question. Participants could not move back and change their previous answers in the survey flow.

## RESULTS

### Data cleaning and retention rate

A total of 23,464 participants clicked on a link and started the PROTECT survey. Upon further inspection of our data set we determined that of these participants only 15,264 participants (65.1% of participants who started the survey) completed 95% or more of the survey. We further cleaned the data set by removing any responses that were completed in less than 300 seconds (about 5 minutes), this reduced the total number of responses to 14,877 (63.4% of participants who started the survey). The median time in which a survey was completed was 22.5 minutes (IQR: 18.0 minutes to 29.6 minutes). Next, responses were reviewed to identify any duplicate responses, this resulted in the removal of further responses, and brought our survey response number to 14,861 (63.3% of participants who started the survey). Responses were then removed from participants who did not indicate their age or that indicated that they were under 18 years of age, which brought our survey response number to 14,823 (63.2% of participants who started the survey). Finally, responses were included into the final data set from MSM and trans* individuals who completed the survey via a version of the survey that was promoted to heterosexual populations in Europe, which brought out final survey response number to 15,458 (see Table 2).

**Table 2:** Total responses to questionnaire

| Country | Data source | Survey data entries | Over 95% complete | Took more than 300 sec to complete | After duplication removed | Excluding underage or no age provided (-38) | MSM included from hetero campaign |
| --- | --- | --- | --- | --- | --- | --- | --- |
| Austria | Dating app | 23 | 16 | 16 | 16 |  |  |
|  | Social media | 299 | 206 | 204 | 204 |  |  |
| Belgium | Dating app | 331 | 240 | 235 | 235 |  |  |
|  | Social media | 388 | 274 | 272 | 272 |  |  |
| Cyprus | Dating app | 51 | 30 | 29 | 29 |  |  |
|  | Social media | 37 | 32 | 32 | 32 |  |  |
| Czech Republic | Dating app | 290 | 178 | 174 | 174 |  |  |
|  | Social media | 314 | 185 | 185 | 185 |  |  |
| Denmark | Dating app | 154 | 105 | 101 | 101 |  |  |
|  | Social media | 141 | 103 | 102 | 102 |  |  |
| Finland | Dating app | 154 | 113 | 109 | 109 |  |  |
|  | Social media | 161 | 116 | 116 | 116 |  |  |
| France | Dating app | 2464 | 1547 | 1490 | 1481 |  |  |
|  | Social media | 965 | 584 | 578 | 578 |  |  |
| Germany | Dating app | 1603 | 1086 | 1057 | 1056 |  |  |
|  | Social media | 1620 | 1135 | 1130 | 1130 |  |  |
| Greece | Dating app | 221 | 128 | 126 | 126 |  |  |
|  | Social media | 52 | 36 | 36 | 36 |  |  |
| Ireland | Dating app | 394 | 266 | 254 | 254 |  |  |
|  | Social media | 261 | 177 | 173 | 173 |  |  |
| Italy | Dating app | 2416 | 1495 | 1439 | 1434 |  |  |
|  | Social media | 702 | 417 | 407 | 407 |  |  |
| Luxemburg | Dating app | 36 | 24 | 24 | 24 |  |  |
|  | Social media | 55 | 31 | 29 | 29 |  |  |
| Netherlands | Dating app | 1130 | 782 | 757 | 757 |  |  |
|  | Social media | 1047 | 697 | 695 | 695 |  |  |
| Norway | Dating app | 101 | 53 | 49 | 49 |  |  |
|  | Social media | 92 | 63 | 61 | 61 |  |  |
| Poland | Dating app | 258 | 165 | 157 | 157 |  |  |
|  | Social media | 214 | 140 | 137 | 137 |  |  |
| Portugal | Dating app | 325 | 219 | 211 | 211 |  |  |
|  | Social media | 381 | 260 | 255 | 255 |  |  |
| Spain | Dating app | 2441 | 1485 | 1431 | 1431 |  |  |
|  | Social media | 1111 | 695 | 680 | 679 |  |  |
| Sweden | Dating app | 292 | 191 | 180 | 180 |  |  |
|  | Social media | 156 | 110 | 107 | 107 |  |  |
| Switzerland | Dating app | 97 | 66 | 65 | 65 |  |  |
|  | Social media | 258 | 177 | 173 | 173 |  |  |
| United Kingdom | Dating app | 1052 | 691 | 675 | 675 |  |  |
|  | Social media | 863 | 565 | 555 | 555 |  |  |
| Flyer |  | 4 | 2 | 2 | 2 |  |  |
| Website |  | 510 | 379 | 369 | 369 |  |  |
| Hetero campaign |  |  |  |  |  |  | 635 |
| Total |  | 23464 | 15264 | 14877 | 14861 | 14823 | 15458 |

### Returns

The survey could be completed via links presented on promotional materials found by participants on social media, dating apps and the project website or flyers. Most participants participated via links promoted via dating apps (57.6%), followed by links promoted via social media (39.9%) and the project website and flyers (2.5%). The proportion of the sample recruited via these promotional sources differed by country (see Table 2). The greatest number of surveys were collected from residents of Germany followed by Spain, France, Italy, the Netherlands and the UK (see Table 3).

**Table 3:** Sample characteristics

| Variable | Answer options | n | % |
| --- | --- | --- | --- |
| Gender identity | Male | 15031 | 97.2 |
|  | Non-binary | 213 | 1.4 |
|  | Trans* male | 85 | 0.5 |
|  | Trans* female | 49 | 0.3 |
|  | Other | 35 | 0.2 |
|  | Prefer not to say | 26 | 0.2 |
|  | Female | 19 | 0.1 |
| Sexual orientation | Homosexual, gay | 12803 | 82.8 |
|  | Bisexual | 1974 | 12.8 |
|  | Pansexual | 230 | 1.5 |
|  | Queer | 216 | 1.4 |
|  | Heterosexual | 92 | 0.6 |
|  | Prefer not to say | 87 | 0.6 |
|  | Other | 56 | 0.4 |
| Age (years) | 18-24 | 1093 | 7.1 |
|  | 25-29 | 1644 | 10.6 |
|  | 30-39 | 4651 | 30.1 |
|  | 40-49 | 4012 | 26 |
|  | 50-59 | 2538 | 16.4 |
|  | 60-69 | 810 | 5.2 |
|  | 70+ | 710 | 4.6 |
| Size of place of residence | A very big city or town (a million or more people) | 4566 | 29.5 |
|  | A big city or town (500,000-999,999 people) | 2480 | 16 |
|  | A medium-sized city or town (100,000-499,999 people) | 3477 | 22.5 |
|  | A small city or town (10,000-99,999 people) | 3026 | 19.6 |
|  | A village / the countryside (less than 10,000 people) | 1909 | 12.3 |
| Housing situation | I live in my own apartment/house | 6841 | 44.3 |
|  | I live in a rented apartment/house | 5287 | 34.2 |
|  | I live with my parents/family members | 1371 | 8.9 |
|  | I rent a room in a shared apartment | 1293 | 8.4 |
|  | I live with my partner (rent-free) | 578 | 3.7 |
|  | I am couch-surfing/staying with different people | 58 | 0.4 |
|  | I am homeless | 30 | 0.2 |
| Migrant | Local | 10521 | 68.1 |
|  | First Generation migrant | 3647 | 23.6 |
|  | Second Generation migrant | 1290 | 8.3 |
| Education level | I do not have a high school diploma | 377 | 2.4 |
|  | Secondary education (high school or equivalent) | 4332 | 28 |
|  | Bachelor degree (university or equivalent) | 4722 | 30.5 |
|  | Master degree (university or equivalent) | 5060 | 32.7 |
|  | PhD / Doctorate | 967 | 6.3 |
| **Employment** | Employed | 11866 | 76.8 |
|  | Student | 1087 | 7 |
|  | Other | 1065 | 6.9 |
|  | Unemployed | 751 | 4.9 |
|  | Retired/Medical leave | 689 | 4.5 |
| **Income in euros (gross annual)** | Less than 10,000 | 1805 | 12.2 |
|  | 10,000 - 19,999 | 2114 | 13.7 |
|  | 20,000 - 29,999 | 2498 | 16.2 |
|  | 30,000 - 39,999 | 2211 | 14.3 |
|  | 40,000 - 49,999 | 1713 | 11.1 |
|  | 50,000 - 59,999 | 1299 | 8.4 |
|  | 60,000 - 69,999 | 980 | 6.3 |
|  | 70,000 - 79,999 | 619 | 4 |
|  | 80,000 - 89,999 | 525 | 3.4 |
|  | 90,000 - 99,999 | 378 | 2.4 |
|  | 100,000 - 149,999 | 770 | 5 |
|  | More than 150,000 | 408 | 2.6 |
| **Comfort on current income** | Really struggling on present income | 1062 | 6.9 |
|  | Struggling on present income | 2142 | 13.9 |
|  | Neither comfortable nor struggling on present income | 5522 | 35.7 |
|  | Living comfortably on present income | 5594 | 36.2 |
|  | Living really comfortably on present income | 1138 | 7.4 |
| **Connection to LGBTIQ communities** | Not at all | 1402 | 9.1 |
|  | A little | 3111 | 20.1 |
|  | Moderately | 4832 | 31.3 |
|  | A lot | 3501 | 22.6 |
|  | A great deal | 1977 | 12.8 |
|  | Didn't receive question | 635 | 4.1 |
| **Relationship status** | Single, not dating | 3140 | 20.3 |
|  | Single, dating | 5355 | 34.6 |
|  | In a relationship | 3278 | 21.2 |
|  | In an open/polyamorous relationship | 3609 | 23.3 |
|  | Widowed, not dating | 18 | 0.1 |
|  | Widowed, dating | 58 | 0.4 |
| **Country of residence** | Austria | 257 | 1.7 |
|  | Belgium | 560 | 3.6 |
|  | Cyprus | 63 | 0.4 |
|  | Czech Republic | 382 | 2.5 |
|  | Denmark | 213 | 1.4 |
|  | Finland | 222 | 1.4 |
|  | France | 2114 | 13.7 |
|  | Germany | 2438 | 15.8 |
|  | Greece | 184 | 1.2 |
|  | Ireland | 424 | 2.7 |
|  | Italy | 1889 | 12.2 |
|  | Luxembourg | 63 | 0.4 |
|  | Netherlands | 1530 | 9.9 |
|  | Norway | 155 | 1 |
|  | Other | 87 | 0.6 |
|  | Poland | 323 | 2.1 |
|  | Portugal | 491 | 3.2 |
|  | Spain | 2185 | 14.1 |
|  | Sweden | 305 | 2 |
|  | Switzerland | 272 | 1.8 |
|  | United Kingdom | 1301 | 8.4 |
| **PrEP use history** | Current | 6512 | 42.1 |
|  | Former | 993 | 6.4 |
|  | Naive | 7953 | 51.4 |
| **Current PrEP regimen** | Daily | 3760 | 24.3 |
|  | Mixed use | 977 | 6.3 |
|  | On-demand | 1775 | 11.5 |
| **Number of sex partners** | 0 | 568 | 3.7 |
|  | 1 | 995 | 6.4 |
|  | 2 - 10 | 5008 | 32.4 |
|  | 10 - 50 | 5482 | 35.5 |
|  | 51-100 | 844 | 5.5 |
|  | 101-150 | 210 | 1.4 |
|  | 150+ | 2351 | 15.2 |
| **Anal sex in past 6 months** | No | 1799 | 11.6 |
|  | Yes | 13659 | 88.4 |
| **Condomless anal intercourse in past 6 months** | No | 2563 | 16.6 |
|  | Yes | 12895 | 83.4 |
| **Substance use in past 6 months to enhance/facilitate sex** | No | 10723 | 69.4 |
|  | Yes | 4735 | 30.6 |
| **Chemsex in past 6 months** | No | 13263 | 85.8 |
|  | Yes | 2195 | 14.2 |
| **Payment given for sex ever** | No | 12478 | 80.7 |
|  | Yes | 2980 | 19.3 |
| **Payment received for sex ever** | No | 12788 | 82.7 |
|  | Yes | 2670 | 17.3 |
| **HIV testing frequency** | Frequently testing | 6700 | 43.3 |
|  | Every six months | 2740 | 17.7 |
|  | Less than once per year | 1742 | 11.3 |
|  | Never | 1114 | 7.2 |
|  | Once per year | 2434 | 15.7 |
|  | NA | 728 | 4.7 |
| **HIV status** | I don't know if I have HIV | 1345 | 8.7 |
|  | No, I do not have HIV | 13385 | 86.6 |
|  | Yes, I have HIV | 728 | 4.7 |
| **STI testing frequency** | Frequently testing | 6532 | 42.3 |
|  | Every six months | 2844 | 18.4 |
|  | Less than once per year | 2192 | 14.2 |
|  | Never | 1587 | 10.3 |
|  | Once per year | 2303 | 14.9 |
| **STI diagnosis ever** | I don't know | 151 | 1 |
|  | No, never | 4381 | 28.3 |
|  | Yes, in the past 12 months | 1746 | 11.3 |
|  | Yes, in the past 6 months | 2704 | 17.5 |
|  | Yes, more than a year ago | 4889 | 31.6 |
|  | Not Applicable | 1587 | 10.3 |
| **STI diagnosis types** | Gonorrhea | 5134 | 33.2 |
|  | Chlamydia | 4681 | 30.3 |
|  | Syphilis | 3319 | 21.5 |
| **Vaccines received** | Hepatitis B | 11477 | 74.2 |
|  | Hepatitis A | 9741 | 63 |
|  | Mpox | 6024 | 39 |
|  | HPV | 3908 | 25.3 |
|  | Covid-19 | 14161 | 91.6 |
|  | Meningococcal | 3360 | 21.7 |
|  | None of the above | 296 | 1.9 |

### Datasets produced

The PROTECT Survey main data set contains 15,458 participants. Country level data sets have been created for all countries that reached over 200 participants, this includes all countries previously listed except Norway and Luxembourg. Greece and Cyprus were both under 200 participants but were combined to create one data set for both countries. The datasets have been analyzed and descriptive data has been prepared and will be shared publicly on the project website (https://protect-study.eu/). Maastricht University, Faculty of Psychology and Neuroscience, owns the datasets and will manage any third-party requests.

### Sample characteristics

Considering all of the MSM/trans* participants, most identified as cis-gendered men (97.2%) with smaller proportions identifying as non-binary (1.4%) or trans* male or female (0.8%) or other (0.5%). Most identified as gay or homosexual (82.8%) with some identifying as bisexual (12.8%) or other (4.4%). The mean age of participants was 40.9 years (SD 11.9; range 18 to 94), and the median was 40.0 years (IQR: 32.0 - 50.0 years). Nearly a third of the sample lived in cities over a million people (29.5%), however many also lived in small towns of between 10,000 and 99,000 people (19.6%) or in villages of under 10,000 people (12.3%). Most participants lived in the country that they and their parents were born in (68.1%) however many first-generation migrants (23.6%) or second-generation migrants (8.3%) participated. The majority of the sample (69.6%) had at least completed a bachelor’s degree at a university or equivalent, or higher. Over three quarters of our sample, (76.8%) were employed and about one fifth of participants (20.8%) reported they were struggling or really struggling to get by on their present income. A moderate to a great deal of connection with local LGBTI communities was reported among most participants (66.7%) in the sample.

More than half of the sample was single (54.9%) with others reporting being in an open or polyamorous relationship (23.3%) and in a monogamous relationship (21.2%). The majority of the sample had 10 or more sexual partners in the past 6 months (57.6%) with more than a fifth of participants having more than 50 (22.1%). Having ever given payment for sex was reported in a substantial minority of the sample (19.3%). Having ever received payment for sex was also reported among a substantial minority of the sample (17.3%).

The majority reported having had anal sex in the past 6 months (88.4%). When asked about their use of oral PrEP, many reported never having taken it (51.4%) with smaller proportions reporting being a current user (42.1%) or a former user (6.4%). Among current oral PrEP users, most reported taking it daily (57.7%) with less taking it on-demand (27.3%) or a mix of periods of daily use and on-demand use (15.0%). Condomless anal intercourse was reported by the majority of participants (83.4%).

Most participants reported testing for HIV at least once per year or more frequently (76.7%) and a minority of the sample reported that they had received a positive diagnosis for HIV (4.7%). Testing for STIs at least once per year or more frequency was also observed among most participants (75.6%) and the majority of participants had ever been diagnosed with an STI (60.4%). The most common STIs diagnosed were Gonorrhea (33.2%), Chlamydia (30.3%) and Syphilis (21.5%). The most common vaccines received for STIs among participants were Hepatitis B (74.2%), Hepatitis A (63.0%) and Mpox (39.0%).

Nearly one third of participants (30.6%) reported using at least one recreational drug in the past six months to facilitate or enhance sexual activity, with less reporting chemsex (14.2%), defined here as use of one or more of the following particular drugs to facilitate sex (methamphetamine, GHB/GBL, mephedrone, ketamine, cocaine).

## DISCUSSION

MSM and trans* people responded well to the opportunity to participate in this survey on potential LA-PrEP use in Europe. The retention rate that we observed with our MSM and trans* component of the survey (63.2%) was comparable to other major European online surveys of this population, such as the EMIS survey retention rate from smartphones in 2017 (27). Smartphones have been shown to be the device from which most online activity is currently taking place amongst people of all ages and demographics in Europe (37). It is likely that most participants of the PROTECT survey completed their responses via their smartphone. The greatest proportion of participants that were excluded from our final data set were ones who did not complete more than 95% of the survey. It is likely that some potential participants were fatigued or did not have the time to fully complete the survey. The median completion time of 22.5 minutes demonstrates that completing the survey required a considerable time commitment.

The sample that was obtained in the PROTECT survey was large, diverse and its sociodemographic variables mirror other large European online surveys of these populations (27). The majority of the sample were cis-gendered men who are gay identifying and are in their thirties and forties. The significant proportion of participants who were not locals and were 1^st^ of 2^nd^ generation migrants (31.9%), may reflect the mobility that citizens of the European Union (EU) possess to reside in any EU country. This high proportion of migrants may also point to the migration patterns of these populations, which have been reported to coalesce in urban centers and which experience greater family breakdown due to their sexual orientation or gender identity and thus more frequent experiences of migration (38-41). Most participants were employed, had at least a bachelors’ degree and were not struggling on their present income, which provides us with a sample that mirrors the experience of many MSM in Europe, and a sample who could potentially pay for a course of LA-PrEP if it was available to them. A third of participants (33.3%) reported not having much of a connection to their local LGBTI communities. This group may be harder to engage with messages about HIV prevention and in particular about LA-PrEP if and when it becomes available.

Our sample was sexually active, with most participants having had anal sex and having had more than 10 sexual partners in the past 6 months. Participants tested often for HIV and STIs indicating that the participants as a group are aware and engaged with their sexual health. The sample provides an interesting, nearly 50/50 split of PrEP naïve versus oral PrEP experienced users (current and former). The considerable proportion of PrEP experienced users is higher than most estimates of current PrEP use among MSM in Europe (42). As the survey was promoted using materials that mentioned PrEP, it may have also attracted more people who had experience with it and are highly sexually active. Over a quarter of the current PrEP users reported using oral PrEP on demand, with a smaller proportion using on demand occasionally interspersed with daily use. This finding demonstrates and reinforces existing understandings of how oral PrEP users in Europe are already using PrEP in alternative ways to the standard daily pill to meet their HIV prevention needs.

Our results indicate that MSM and trans* individuals can be engaged effectively using promotional material over both social media and dating apps. As has been shown in other major online surveys of MSM, places where MSM are looking for sex and relationships can be effectively used to engage MSM on topics of sexual health (27). The larger countries in our study, such as the EU-5 (Germany, France, Italy, Spain and the UK) have higher proportions of their samples responding via dating apps. This could be associated with larger concentrations of MSM and trans people located in the major European urban centers located in these countries (38, 39). Countries with smaller populations, such as Austria, Norway and Switzerland for example, were more challenging to engage over dating apps. Over social media MSM and trans people are also active and can be engaged, however as our data indicates the numbers of surveys generated via social media also varies considerably by country (see table 2). This could be related to how MSM and trans* people are using social media to connect, and the migration and resettlement patterns observed among MSM and trans populations in Europe. Further research into which platforms are more active in each country and on how these populations respond to different types of marketing materials could help future efforts to engage these populations in research.

Obtaining a large sample for online surveys is becoming increasingly difficult. With ever shortening attention spans and increasing concerns over data privacy the desire to participate in surveys may be dwindling (43-45). Furthermore, MSM and trans people often experience stigma and discrimination about their sexual behaviors and may feel wariness to participate in research due to lack of trust or perceived negative consequences. In light of these factors the results obtained in the PROTECT survey are encouraging. Challenges were experienced with data collection in some countries and may reflect how and where MSM and trans* individuals find and connect with each other.

This research has a number of limitations. The dating apps do not have precisely the same memberships across the twenty countries in which the survey was conducted. This led to different numbers of surveys obtained in different locations based on those platforms. The number and distribution of members that they can display an advertisement to in a country at a specific time limit the potential reach of the advertisements on gay dating apps. The survey did take about 20 to 25 minutes to complete and as such, people who have less time to give to this kind of activity did not complete the survey and were not included in the sample. This group may have been less privileged and would be interesting to be better understood. Shorter and perhaps more targeted surveys could be employed to engage people who may not have the time to complete a survey as lengthy as PROTECT. Our survey did not provide incentives or compensation for participation, by providing this in future perhaps some of this time-restricted group may have been retained.

## CONCLUSION

MSM and trans* individuals across Europe were effectively engaged by tailored promotional material to participate in an online survey on their sexual health. Participants were obtained from twenty countries and via promotional material both on social media apps and dating apps. Greater numbers of participants were obtained in larger European countries with perhaps larger communities of MSM and trans people. The sample obtained is representative of these communities across Europe and provides opportunities to explore HIV prevention behavior and beliefs, and in particular interest and intention to use LA-PrEP.

## Author’s contribution statement

**JK, H**W, and KJJ contributed to the analysis study concept and design. JK, HW, HZ, and KJJ contributed to data collection. JK, HW and KJJ contributed to data analyses and/or interpretation of the data. JK drafted the manuscript. All authors critically revised the manuscript. All authors approved the final version for publication.

## Ethics statement

The Ethics Review Committee Psychology and Neuroscience of Maastricht University granted ethical approval for the study (OZL_262_08_01_2023_S21)

## Data sharing statement

Maastricht University, Faculty of Psychology and Neuroscience, owns the datasets and will manage any third-party requests.

## SUPPLEMENTARY MATERIALS

In addition to the scales presented in the Methods section of this manuscript, the scales described below were also included in the PROTECT survey.

### Comfort on current income

Comfort on current income was assessed using a 1-item scale that participants responded to by selecting a response on a 5-point Likert scale from 1 (really struggling on present income) to 5 (living really comfortably on present income). Participants were asked to rate the following item, “*Which of these phrases would you say comes closest to your feelings about your income these days?*”

### Connection to LGBTQI+ communities scale

Connection to LGBTQI+ communities was assessed using a 1-item scale that participants responded to by selecting a response on a 5-point Likert scale from 1 (not at all) to 5 (a great deal). Participants were asked to rate the following item, “*How connected do you feel to Lesbian, Gay, Bisexual, Transgender, Queer or Intersex (LGBTIQ+) communities?*”

### Position taken during anal sex scale

Position most often taken during anal sex was assessed using a 1-item scale that participants responded to by selecting a response on a 5-point Likert scale from 1 (always top) to 5 (always bottom), an additional response option of (I don’t have anal sex) was also included. Participants were asked to rate the following item, “*What position (receptive/bottom and/or insertive/top) are you most likely to take when having anal sex?*”

### Reasons for stopping oral PrEP

Reasons for stopping oral PrEP, among former PrEP using participants, were assessed using a 15-item scale that participants responded to by selecting a response on a 5-point Likert scale from 1 (strongly disagree) to 5 (strongly agree). For example, participants were asked to rate the following item, “*I don’t want to take medication every day”.* The Cronbach’s alpha for this scale was good (α = 0.76).

### Likelihood of oral PrEP continuation scale

The likelihood of continuing to use oral PrEP over the next two years was assessed using a 1-item scale that participants responded to by selecting a response on a 5-point Likert scale from 1 (Extremely unlikely) to 5 (Extremely likely). Participants were asked to rate the following item, “*How likely is it that you are going to use oral PrEP for the next two years?*”

### Likelihood of shortening intake window for on demand PrEP before sex

The likelihood of shortening intake window for on demand PrEP before sex was assessed using a 1-item scale that participants responded to by selecting a response on a 5-point Likert scale from 1 (Never) to 5 (Always). Participants were asked to rate the following item, “*Thinking about the 2 pills for oral PrEP on demand that you must take at least 2 hours before sex, how often do you shorten this period?*”

### Likelihood of forgetting to take PrEP after sex when using PrEP on demand scale

The likelihood of forgetting to take PrEP after sex when using PrEP on demand was assessed using a 1-item scale that participants responded to by selecting a response on a 5-point Likert scale from 1 (Never) to 5 (Always). Participants were asked to rate the following item, “*Thinking about the two pills for oral PrEP on demand that you must take after sexual activity (24 and 48 hours after the initial two pill dose), how often do you forget to take one or both of these pills?”*

### Ease of obtaining PrEP scale

The ease of obtaining oral PrEP, was assessed using a 9-item scale that participants responded to by selecting a response on a 5-point Likert scale from 1 (strongly disagree) to 5 (strongly agree). For example, participants were asked to rate the following item, “*PrEP is included in my health insurance”.* The Cronbach’s alpha for this scale was acceptable (α = 0.61).

### Barriers to obtaining oral PrEP

Barriers to obtaining oral PrEP, were assessed using a 4-item scale that participants responded to by selecting a response on a 5-point Likert scale from 1 (strongly disagree) to 5 (strongly agree) an additional response option of (does not apply) was also included. For example, participants were asked to rate the following item, “*I could not use oral PrEP because it was too costly for me (either the pills, or the tests associated with PrEP use)”.* The Cronbach’s alpha for this scale was very good (α = 0.90).

### Willingness to pay for LA PrEP assesment

Willingness to pay for LA PrEP was assessed with 3-items describing different reimbursement scenarios that participants responded to by selecting a response on a 5-point Likert scale from 1 (extremely unwilling) to 5 (extremely willing). For example, participants were asked to rate the following item, “*In each of the following scenarios, to what extent would you be willing to pay towards your access of LA PrEP?* – 1. *LA PrEP is officially authorised in my country but at a high cost.”*

### Price willing to pay for LA PrEP scale

The price participant were willing to pay for LA PrEP was assessed using a 6-item scale that participants responded to by selecting a response on a 5-point Likert scale from 1 (extremely unlikely) to 5 (extremely likely). For example, participants were asked to rate the following item, “*You have indicated that you are willing to use LA PrEP when you can afford it without it being included in your health insurance. Can you let us know at which price level you are likely willing to pay for it?* – 1. *Under 50 euros per month.”* The Cronbach’s alpha for this scale was good (α = 0.73).

### Worries with LA PrEP use

Worries with LA PrEP use was assessed using a 9-items that participants responded to by selecting a response on a 5-point Likert scale from 1 (strongly disagree) to 5 (strongly agree). For example, participants were asked to rate the following item, “*I have a fear of needles”*.

### Fear of PrEP interacting with hormones

Among trans* participants, fear of PrEP interacting with hormone use was assessed using a 1-item scale that participants responded to by selecting a response on a 5-point Likert scale from 1 (a great deal) to 5 (none at all) an additional response option of (I do not use hormones) was also included. Participants were asked to rate the following item, “*Are you afraid of PrEP negatively interacting with your hormone use?”*

### Worry of LA PrEP affecting bone density

Among trans* participants, worry of PrEP affecting bone density was assessed using a 1-item scale that participants responded to by selecting a response on a 5-point Likert scale from 1 (a great deal) to 5 (none at all). Participants were asked to rate the following item, “*Are you worries that LA PrEP will negatively impact your bone mineral density?”*

### Likelihood of making it to appointment every 2 months for LA PrEP

The likelihood of making it to an appointment every 2 months for LA PrEP was assessed using a 1-item scale that participants responded to by selecting a response on a 5-point Likert scale from 1 (extremely unlikely) to 5 (extremely likely). Participants were asked to rate the following item, “*You have indicated that you are willing to start using LA PrEP every 2 months. Imagine you were to use LA PrEP, how likely would you be able to make it to the appointment to receive the injection?”*

### Likelihood of combining oral & LA PrEP

The likelihood of combining oral and LA PrEP was assessed using a 1-item scale that participants responded to by selecting a response on a 5-point Likert scale from 1 (extremely unlikely) to 5 (extremely likely). Participants were asked to rate the following item, “*How likely are you interested in combining oral PrEP and LA PrEP regimens one after the other? We refer to interchangeable use, for example a period of LA PrEP use followed by oral PrEP use.”*

### Likelihood of combining oral &LA PrEP at particular times of the year

The likelihood of combining oral and LA PrEP during particular times of the year was assessed using a 1-item scale that participants responded to by selecting a response on a 5-point Likert scale from 1 (extremely unlikely) to 5 (extremely likely). Participants were asked to rate the following item, “*How likely are you interested in combining oral PrEP and LA PrEP regimens at different times of the year (e.g. using mainly oral PrEP and then using LA PrEP when you know you will have more frequent sex)?”*

### Likelihood of travelling for access to LA PrEP

The likelihood of travelling for access to LA PrEP was assessed using a 1-item scale that participants responded to by selecting a response on a 5-point Likert scale from 1 (extremely unlikely) to 5 (extremely likely). Participants were asked to rate the following item, “*If you travel to a country where LA PrEP is available and affordable, how likely are you to try to use it there?”*

### Worry about transmitting HIV to partners

Among participants living with HIV, worry about transmitting HIV to partners was assessed using a 1-item scale that participants responded to by selecting a response on a 5-point Likert scale from 1 (never) to 5 (always). Participants were asked to rate the following item, “*How often are you worried that you will pass on HIV to your sexual partners?”*

### Preference for partners to use LA PrEP

Among participants living with HIV, preference for partners to use LA PrEP was assessed using a 1-item scale that participants responded to by selecting a response on a 5-point Likert scale from 1 (definitely yes) to 5 (definitely not). Participants were asked to rate the following item, “*After learning (more) about LA PrEP, would you prefer that your partner uses LA PrEP, instead of oral PrEP, if it is available in your country?”*

### Willingness to pay for partners’ LA PrEP

Among participants living with HIV, willingness to pay for their partners LA PrEP was assessed using a 1-item scale that participants responded to by selecting a response on a 5-point Likert scale from 1 (definitely yes) to 5 (definitely not). Participants were asked to rate the following item, “*Would you be willing to help pay for your partner to take LA PrEP?”*

### Comfort with missing own HIV treatment if partner uses LA PrEP

Among participants living with HIV, comfort with missing their own HIV treatment if their partner is on LA PrEP was assessed using a 1-item scale that participants responded to by selecting a response on a 5-point Likert scale from 1 (not at all comfortable) to 5 (comfortable to miss a dose). Participants were asked to rate the following item, “*Knowing that your partner is taking LA PrEP, how comfortable would you feel about missing doses of your own antiretroviral regimen?”*

### Comfort with missing own HIV treatment if partner uses oral PrEP

Among participants living with HIV, comfort with missing their own HIV treatment if their partner is on oral PrEP was assessed using a 1-item scale that participants responded to by selecting a response on a 5-point Likert scale from 1 (not at all comfortable) to 5 (comfortable to miss a dose). Participants were asked to rate the following item, “*Knowing that your partner is taking oral PrEP, how comfortable would you feel about missing doses of your own antiretroviral regimen?”*

### Attitudes and beliefs towards LA PrEP and LA ART

Among participants living with HIV, attitudes and beliefs towards LA PrEP and long-acting antiretroviral therapy (LA ART) were assessed using 4-items that participants responded to by selecting a response on a 5-point Likert scale from 1 (strongly disagree) to 5 (strongly agree). For example, participants were asked to rate the following item, “*LA PrEP has been proven to be effective at preventing the transmission of HIV to HIV negative people.”*

